# Automated Brain Segmentation in Accelerated T2-Weighted MRI: Effects of Deep Learning-Based Reconstruction

**DOI:** 10.64898/2026.09.08.26362537

**Authors:** Julia Lasek, Rafał Obuchowicz, Michał Strzelecki, Ireneusz Dwojak, Wojciech Kanas, Izabella Obuchowicz, Artur Krzyżak, Jarosław Minkowski

## Abstract

**Background:** Automated brain volumetry is increasingly used for clinical and research assessment, but its outputs depend on acquisition and reconstruction settings. We evaluated how parallel-imaging acceleration and vendor deep-learning reconstruction (Siemens Deep Resolve) affect automated volumetry on T2-weighted turbo spin-echo MRI, and whether deep-learning reconstruction stabilises measurements across protocols.

**Methods:** Four healthy volunteers each underwent 14 T2-TSE-TRA acquisitions on a 1.5 T scanner, covering seven protocols (baseline, GRAPPA R = 2, 3, 4, and SMS ×2, ×3, ×4), with each protocol acquired separately once with Deep Resolve off and once with Deep Resolve on, for 56 acquisitions in total. Six pipelines were applied: SynthSeg, OpenMAP-T2, and GOUHFI 2.0 for subcortical parcellation, and TotalSegmentator MRI, HD-BET, and NV-Segment-CTMR for whole-brain masking. A three-rater STAPLE consensus of six subcortical structures on the baseline acquisition served as the expert reference. Reference-relative accuracy (absolute percentage error), cross-protocol dispersion (coefficient of variation), and spatial reproducibility (Dice coefficient and 95th-percentile surface distance) were compared between reconstruction states.

**Results:** Deep Resolve improved reference-relative accuracy most for GOUHFI 2.0 (pooled median absolute percentage error 28.4% to 21.9%), modestly for SynthSeg (16.2% to 15.5%), and had a mixed, structure-dependent effect for OpenMAP-T2. It approximately halved cross-protocol dispersion for all three parcellation tools (coefficient of variation 2.62% to 1.39% for SynthSeg, 4.42% to 2.14% for OpenMAP-T2, and 21.75% to 7.87% for GOUHFI) and improved spatial reproducibility in parallel. It eliminated the systematic GOUHFI under-segmentation seen at high acceleration under conventional reconstruction (median deviation −31.6% at GRAPPA R = 4; 14 failure events under conventional reconstruction, none with Deep Resolve). Whole-brain masking was robust in both states.

**Conclusions:** Deep Resolve improved the robustness of automated brain segmentation in accelerated T2-weighted MRI, reducing cross-protocol variability and improving spatial reproducibility, with the largest reduction in reference-relative error in the most acceleration-sensitive pipeline. The elimination of severe segmentation failures observed under conventional reconstruction highlights its potential to support reliable volumetry at higher acceleration. Reconstruction optimization therefore offers a practical route to more consistent quantitative measurements across acquisition protocols.

## 1. Introduction

Quantitative volumetric MRI underpins a growing range of clinical decisions. In neurodegenerative disease, regional atrophy, particularly of the hippocampus and of the whole-brain parenchyma, serves as an imaging biomarker for diagnosis, staging, and monitoring of treatment response, and increasingly informs eligibility and follow-up in disease-modifying therapy [1]. In demyelinating disease, the brain parenchymal fraction and ventricular enlargement track neurodegeneration and long-term disability independently of focal lesion burden [2]. Ventricular volume is likewise central to the assessment of hydrocephalus and of atrophic ventriculomegaly [3]. What these applications share is a reliance not on a single measurement but on small longitudinal or cross-sectional differences: expected annualised atrophy rates lie in the low single-digit percentage range, comparable in magnitude to the measurement variability that acquisition and reconstruction changes can themselves introduce. Measurement stability across protocols is therefore not a technical nicety but a precondition for clinical interpretability.

Quantitative volumetric analysis of MRI-derived brain structures is central to the assessment of neurodegenerative disease, of demyelinating disorders, and of longitudinal treatment response. Manual delineation by expert raters is time-consuming, requires specialised expertise, and remains susceptible to inter-rater variability, limiting its feasibility in large imaging datasets [4]. Automated segmentation pipelines provide a scalable alternative to manual delineation and are increasingly used for research and clinical volumetric assessment. The clinical value of their output, in particular hippocampal, ventricular, and whole-brain volumes, depends critically on the stability of the derived measurements across acquisition and reconstruction conditions.

Contemporary clinical MRI increasingly relies on parallel imaging acceleration to reduce examination time. Two widely used acceleration approaches are GRAPPA and simultaneous multi-slice imaging. GRAPPA reconstructs missing k-space samples using autocalibration data and the spatial encoding provided by multiple receiver coils [5]. Simultaneous multi-slice (SMS) or multiband imaging excites multiple slices in a single radio-frequency pulse and separates them at reconstruction [6, 7]. Both techniques can alter effective SNR and artefact structure, particularly at higher acceleration factors, through g-factor-related noise amplification, imperfect slice separation, and residual aliasing [7]. These effects propagate into downstream quantitative measurements. Dieckmeyer et al. [8] demonstrated acceleration-dependent, software-specific biases in brain tissue volumes derived from compressed-SENSE-accelerated 3D T1-weighted imaging. A related effect has been documented in lesion volumetry, where Ghankot et al. [9] showed that voxel-size changes alter the measured volume of vestibular schwannomas independently of the underlying lesion. Automated volumetry is therefore not invariant to protocol changes, and any strategy that preserves downstream measurement fidelity under aggressive acceleration is of direct clinical interest.

Vendor-specific deep-learning image reconstruction pipelines have been introduced as one such strategy. Siemens Deep Resolve comprises a family of deep-learning-based reconstruction and image-enhancement methods designed to support accelerated acquisition through operations such as targeted denoising, resolution enhancement, and reconstruction of undersampled data. Comparable deep-learning reconstruction pipelines are also available from other MRI manufacturers under different trade names, though the present study is restricted to Siemens Deep Resolve. Previous studies have generally reported improved perceived sharpness, noise characteristics, and anatomical delineation after deep-learning reconstruction, although reconstruction-specific artefacts may persist, become more conspicuous, or occasionally mimic pathology [10–13]. Quantitative evidence supports partial preservation of downstream measurements: Bash et al. [14] reported that a 60% reduction in acquisition time achieved with deep-learning reconstruction preserved qualitative image quality and volumetric quantification performance in T1-weighted brain MRI, and Jung et al. [15] reported that deep-learning-reconstructed accelerated 3D T1-weighted acquisitions produced brain-volume estimates whose intraclass agreement with conventional acquisitions approached unity for most cortical structures. Beyond volumetry, the same reconstruction operations reshape voxel-level image texture: on comparable GRAPPA- and SMS-accelerated acquisitions from the same scanner platform, Deep Resolve has been shown to alter the stability of radiomic texture features in a tissue- and acceleration-dependent manner [16].

The generalisability of these observations to downstream segmentation on T2-weighted acquisitions, widely used in clinical practice for lesion characterisation and morphometry, has not been systematically established. Liu et al. [12] evaluated deep-learning reconstruction for T2-weighted turbo spin-echo imaging but focused on diagnostic image quality rather than automated segmentation. The joint effect of parallel-imaging acceleration and Deep Resolve reconstruction on multiple state-of-the-art segmentation pipelines applied to the same prospectively acquired T2-TSE dataset has not, to our knowledge, been reported.

Automated brain segmentation has advanced rapidly. Contrast-agnostic parcellation, exemplified by SynthSeg [17], enables segmentation across MR contrasts without retraining. OpenMAP-T1[18] provides fine-grained parcellation of 280 anatomical regions on T1-weighted images, and its OpenMAP-T2 variant extends the design to T2-weighted acquisitions. GOUHFI 2.0 [19] targets ultra-high-field brain segmentation through a two-stage subcortical and cortical pipeline. Whole-brain masking has migrated from atlas-based methods to deep networks: HD-BET [20] uses a multi-sequence CNN trained for robust skull stripping across contrasts, TotalSegmentator MRI [21] provides sequence-independent segmentation of multiple anatomical structures including a whole-brain compartment, and foundation-model approaches such as VISTA3D [22], support automated segmentation across a wide range of medical imaging tasks.

The primary question of the present study is how sensitive automated volumetric measurements are to acceleration protocol and Deep Resolve reconstruction state on prospectively acquired T2-TSE MRI. Four healthy volunteers were scanned with 14 T2-TSE-TRA protocols per subject (seven acquisition methods, baseline plus GRAPPA R = 2, 3, 4 and SMS ×2, ×3, ×4, each combined with two Deep Resolve states). Six automated segmentation pipelines were applied: SynthSeg [17], OpenMAP-T2 [18], and GOUHFI 2.0 [19] for detailed subcortical parcellation, and TotalSegmentator MRI [21], HD-BET [20], and NV-Segment-CTMR [22] for whole-brain masking. Three independent raters provided manual segmentations of six subcortical structures on the baseline acquisition of each subject.

Accordingly, this study investigated whether MRI acceleration and Deep Resolve reconstruction affect automated measurements of regional and whole-brain volumes. Particular attention was given to whether the observed differences depended on the anatomical structure, segmentation tool, or acceleration protocol. We also examined whether Deep Resolve improved the consistency of measurements obtained from different acquisition settings.

## 2. Methods

### 2.1 Subjects

Four healthy adult volunteers were scanned in a single session each. The study protocol was designed in accordance with guidelines of the Declaration of Helsinki and the Good Clinical Practice Declaration Statement. Written approval was obtained from the Bioethics Committee to conduct this study (No. 11/KBL/OIL/2025 dated 11 March, 2025 and 03 March 2026).

### 2.2 MRI acquisition

All acquisitions were performed on a Siemens MAGNETOM Altea 1.5 T scanner (Siemens Healthineers, Erlangen, Germany) using the vendor head coil. Each subject underwent 14 T2-weighted turbo spin-echo transverse (T2-TSE-TRA) acquisitions covering seven protocols in both Deep Resolve reconstruction states, for a total of 56 acquisitions across the four subjects.

The seven protocols comprised a baseline acquisition without parallel-imaging acceleration, three GRAPPA-accelerated acquisitions (R = 2, 3, 4), and three simultaneous multi-slice acquisitions (SMS ×2, ×3, ×4). The following sequence parameters were used: acquired in-plane resolution 0.513 × 0.513 mm, matrix 364 × 448, slice thickness 6.0 mm, 26 slices, echo train length 4.

Each protocol was acquired twice, once with Deep Resolve turned off and once with Deep Resolve turned on. In the Deep Resolve on state, two vendor modules were active: Gain, an adaptive denoising module that reduces image noise while preserving edges, and Sharp, a super-resolution module that upsamples the reconstructed matrix by a factor of two along both in-plane axes. The reconstructed matrix in the Deep Resolve on state was therefore 728 × 896 with a nominal reconstructed in-plane resolution of 0.257 × 0.257 mm.

### 2.3 Manual segmentation and consensus reference

Three raters independently segmented six subcortical structures on the baseline DR-off acquisition of each subject using 3D Slicer (v5.11). The rater group consisted of two radiologists with 20 and 8 years of experience, respectively, and one medical student. The six structures were: hippocampus, amygdala, caudate nucleus, lateral ventricle, brain stem, and putamen. Bilateral structures were segmented as a single label combining both hemispheres.

A per-structure binary consensus was computed with the Simultaneous Truth and Performance Level Estimation (STAPLE) algorithm [23] as implemented in SimpleITK, using a threshold of 0.5 on the STAPLE foreground probability. STAPLE consensus masks were assembled into per-subject multi-label reference volumes; consensus volumes *V_ref* used in subsequent analyses were computed as the voxel count of the consensus mask multiplied by the acquisition voxel volume.

### 2.4 Automated segmentation pipelines

Six automated segmentation pipelines were applied to every one of the 56 T2-TSE acquisitions:

- SynthSeg [17] - a contrast- and resolution-agnostic deep-learning framework for whole-brain segmentation and labelling of subcortical structures, cortical regions, cerebellum, ventricles, and major tissue compartments
- OpenMAP-T2 - the official T2-weighted implementation of OpenMAP, providing whole-brain parcellation into 280 anatomical regions based on the JHU atlas [18].
- GOUHFI 2.0 [19] - contrast- and resolution-agnostic brain segmentation toolbox comprising two independently trained 3D U-Net models for whole-brain segmentation and cortical parcellation; their output label maps were merged for the present analysis.
- TotalSegmentator MRI [21] - a sequence-independent nnU-Net-based model for multi-anatomical MRI segmentation; only the brain label was retained for the present analysis.
- HD-BET [20] - a dedicated multisequence MRI brain-extraction network producing a binary brain mask.
- NV-Segment-CTMR - a foundation-model segmentation pipeline based on VISTA3D [22]; only the brain label was retained.

### 2.5 Label harmonisation

To enable cross-tool comparison at the level of anatomical structures, tool-specific label identifiers were mapped through an explicit lookup table to 19 regional anatomical groups plus a whole-brain compartment. Additional entries in the mapping are technical placeholders for labels lacking a FreeSurfer equivalent and were excluded from every cross-tool comparison. Left- and right-hemisphere labels within a tool were merged into a single group per structure. Whole-brain masks produced by TotalSegmentator, HD-BET, and NV-Segment-CTMR were mapped to the single group whole-brain. Anatomical volumes were computed as the number of voxels assigned to the group multiplied by the acquisition voxel volume.

### 2.6 Analytic framework

For each subject, acquisition protocol, structure covered by the manual reference (n = 6), and parcellation tool (SynthSeg, OpenMAP-T2, GOUHFI 2.0), the absolute percentage volume error against the STAPLE consensus was computed as

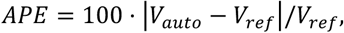

where *V_auto* is the tool-derived volume of the structure on the acquisition and *V_ref* is the STAPLE consensus volume of the structure for that subject, treated as the anatomical volume of the structure and held constant across the acquisitions of a given session. Two summaries of APE were reported: the pooled median within each Deep Resolve state, and the paired within-pair change

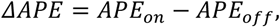

computed within each subject × acquisition × structure pair and then summarised as a paired median. Spatial overlap against the STAPLE consensus was reported only for the baseline DR-off acquisition, using Dice and the 95th-percentile Hausdorff distance.

For every (subject, region of interest, tool, Deep Resolve state) unit, the coefficient of variation of the tool-derived volume across the seven acquisition protocols was computed as

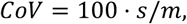

where s is the sample standard deviation and m is the sample mean of the seven per-protocol volumes. Trajectories with fewer than seven scans, attributable to a pipeline failing to produce the label in one of the acquisitions, were excluded from the CoV analysis.

To isolate the effect of acceleration from that of Deep Resolve, deviations from the baseline acquisition of the matching Deep Resolve state were computed as

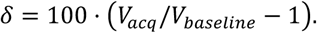

Deviations were reported at the two maximum-acceleration protocols (GRAPPA R = 4 and SMS ×4). Segmentation failures were defined as

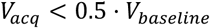

corresponding to a volume output below half of the same-Deep Resolve baseline value.

Paired volume differences between DR-off and DR-on reconstructions of the same nominal protocol were computed as

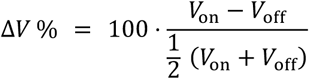

for each subject, structure, and tool, and summarised per acquisition protocol.

Spatial reproducibility of the parcellations across protocols was assessed with the Dice similarity coefficient and the 95th-percentile symmetric surface distance (HD95, in millimetres). For each subject, manual-reference structure, parcellation tool, and Deep Resolve state, two complementary summaries were computed. The mean Dice and HD95 over all pairs of the seven acquisition protocols quantified the overall cross-protocol spatial reproducibility of the segmented structure. In addition, the Dice and HD95 between each accelerated acquisition and the baseline acquisition of the matching Deep Resolve state quantified the change in the spatial extent of the structure with increasing acceleration. Both summaries were reported as medians over subjects and structures within each tool and Deep Resolve state.

### 2.7 Software

Preprocessing, analysis, and figure generation were performed in Python 3.11.9 (Python Software Foundation) using pandas (v2.2.3), NumPy (v1.24.4), SimpleITK (v2.4.1), nibabel (v5.2.1), and matplotlib (v3.10.8). DICOM to NIfTI conversion was performed with dcm2niix.

## 3. Results

All 56 T2-TSE acquisitions were processed with six automated segmentation pipelines. Two questions structured the analysis: whether Deep Resolve reconstruction moves tool-derived volumes closer to the expert reference, and whether it reduces the dispersion of those volumes across acquisition protocols and prevents severe deviations at high acceleration. Aggregate statistics are reported both pooled and, where relevant, at the subject level.

### 3.1 Inter-rater agreement of the expert reference

Median pairwise Dice ranged from 0.86 (putamen) to 0.96 (brain stem) across the 72 rater pairs (Table 1). Agreement was highest for the brain stem and hippocampus and lowest for the putamen. STAPLE consensus segmentations of the four baseline (DR-off, no acceleration) acquisitions were used as the expert reference for all subsequent accuracy analyses.

**Table 1.** Inter-rater agreement (12 pairs per structure).

| Structure | Pairwise Dice, median [IQR] | Range |
| --- | --- | --- |
| Brain stem | 0.96 [0.95, 0.97] | [0.933, 0.979] |
| Hippocampus | 0.93 [0.87, 0.94] | [0.809, 0.966] |
| Amygdala | 0.92 [0.85, 0.93] | [0.762, 0.937] |
| Caudate | 0.90 [0.88, 0.91] | [0.780, 0.920] |
| Lateral ventricle | 0.87 [0.86, 0.90] | [0.828, 0.944] |
| Putamen | 0.86 [0.83, 0.89] | [0.764, 0.942] |

### 3.2 Deep Resolve effect on reference-relative volumetric accuracy

For each subject, acquisition protocol, structure, and parcellation tool, the absolute percentage error (APE) of the automated volume relative to the STAPLE consensus was compared between DR-off and DR-on reconstructions of the same nominal protocol.

The median APE, computed independently for the Deep Resolve off (DR-off) and Deep Resolve on (DR-on) pools, provides a descriptive summary of reference-relative volumetric error in each condition. The paired median ΔAPE was calculated within each subject × acquisition × structure pair as the difference between APE in the DR-on condition and APE in the DR-off condition (APE_DR-on − APE_DR-off) and then summarised across pairs. This measure isolates the within-pair effect of Deep Resolve.

For GOUHFI 2.0, Deep Resolve produced the largest accuracy gain. Pooled median APE decreased from 28.39% to 21.89%, paired median ΔAPE was −3.32 percentage points (pp), and 114 of 168 pairs improved (Table 2). Structure-level effects were dominated by the lateral ventricle (paired median ΔAPE −8.35 pp, 27/28 pairs improved) and the caudate (−6.85 pp, 19/28), with smaller improvements for the brain stem (−3.07 pp) and hippocampus (−2.38 pp). All four subject-level medians were negative (range −7.14 to −0.84 pp).

**Table 2.** Reference-relative volumetric accuracy per structure and tool. APE, absolute percentage error against STAPLE consensus, median; ΔAPE = APE_on - APE_off, paired median (pp).

| Structure | SynthSeg |  |  | OpenMAP-T2 |  |  | GOUHFI |  |  |
| --- | --- | --- | --- | --- | --- | --- | --- | --- | --- |
| | APE<br>DR-off | APE<br>DR-on | ΔAPE | APE<br>DR-off | APE<br>DR-on | ΔAPE | APE<br>DR-off | APE<br>DR-on | ΔAPE |
| Brain stem | 15.66<br>[14.32,<br>16.44] | 15.28<br>[13.45,<br>16.17] | -0.26 | 49.62<br>[47.23,<br>50.90] | 49.98<br>[47.83,<br>51.17] | +0.32 | 40.19<br>[31.05,<br>53.24] | 39.60<br>[27.65,<br>47.44] | -3.07 |
| Hippocampus | 28.09<br>[26.27,<br>37.32] | 27.41<br>[25.17,<br>32.24] | -0.89 | 16.89<br>[15.18,<br>22.38] | 19.63<br>[18.67,<br>27.31] | +1.93 | 27.04<br>[5.53,<br>37.22] | 20.37<br>[4.67,<br>34.09] | -2.38 |
| Amygdala | 4.89<br>[1.99,<br>19.01] | 2.71<br>[1.26,<br>11.05] | -0.42 | 12.74<br>[5.96,<br>30.26] | 13.67<br>[6.00,<br>25.30] | +0.78 | 29.56<br>[14.50,<br>42.37] | 19.58<br>[7.55,<br>37.91] | -0.87 |
| Caudate | 12.67<br>[7.95,<br>18.52] | 11.42<br>[8.87,<br>14.92] | -0.43 | 12.58<br>[6.55,<br>16.26] | 12.93<br>[8.38,<br>21.26] | +1.32 | 24.21<br>[13.76,<br>29.86] | 20.77<br>[12.47,<br>24.33] | -6.85 |
| Lateral<br>ventricle | 37.93<br>[35.13,<br>46.89] | 38.78<br>[35.30,<br>44.30] | -1.00 | 18.36<br>[15.57,<br>20.71] | 13.87<br>[12.86,<br>15.01] | -3.20 | 35.49<br>[28.43,<br>44.83] | 22.44<br>[19.60,<br>35.63] | -8.35 |
| Putamen | 9.70<br>[4.93,<br>12.01] | 10.38<br>[6.73,<br>12.46] | -0.09 | 8.03<br>[4.83,<br>10.18] | 10.43<br>[7.82,<br>12.98] | +2.53 | 12.85<br>[9.45,<br>19.14] | 12.51<br>[8.77,<br>16.13] | -1.73 |
| Pooled | 16.19<br>[9.88,<br>33.02] | 15.53<br>[9.54,<br>28.50] | -0.55 | 16.16<br>[9.36,<br>28.24] | 15.97<br>[11.48,<br>27.31] | +0.52 | 28.39<br>[15.41,<br>40.54] | 21.89<br>[12.53,<br>31.90] | -3.32 |

For SynthSeg, the accuracy gain was small but consistent in direction. Pooled median APE decreased from 16.19% to 15.53%, paired median ΔAPE was −0.55 pp, and 109 of 168 pairs improved. All four subject-level medians were negative (range −1.06 to −0.06 pp).

For OpenMAP-T2, the effect was mixed and structure dependent. Pooled median APE was essentially unchanged (16.16% vs 15.97%) and paired median ΔAPE was +0.52 pp, with 74 of 168 pairs improved. The lateral ventricle improved substantially (paired median ΔAPE −3.20 pp, 25/28 pairs improved) but hippocampus, caudate, and putamen shifted away from the reference (paired median ΔAPE +1.32 to +2.53 pp). Only one of four subject-level medians was negative (range −0.27 to +1.49 pp).

**Figure 1.**
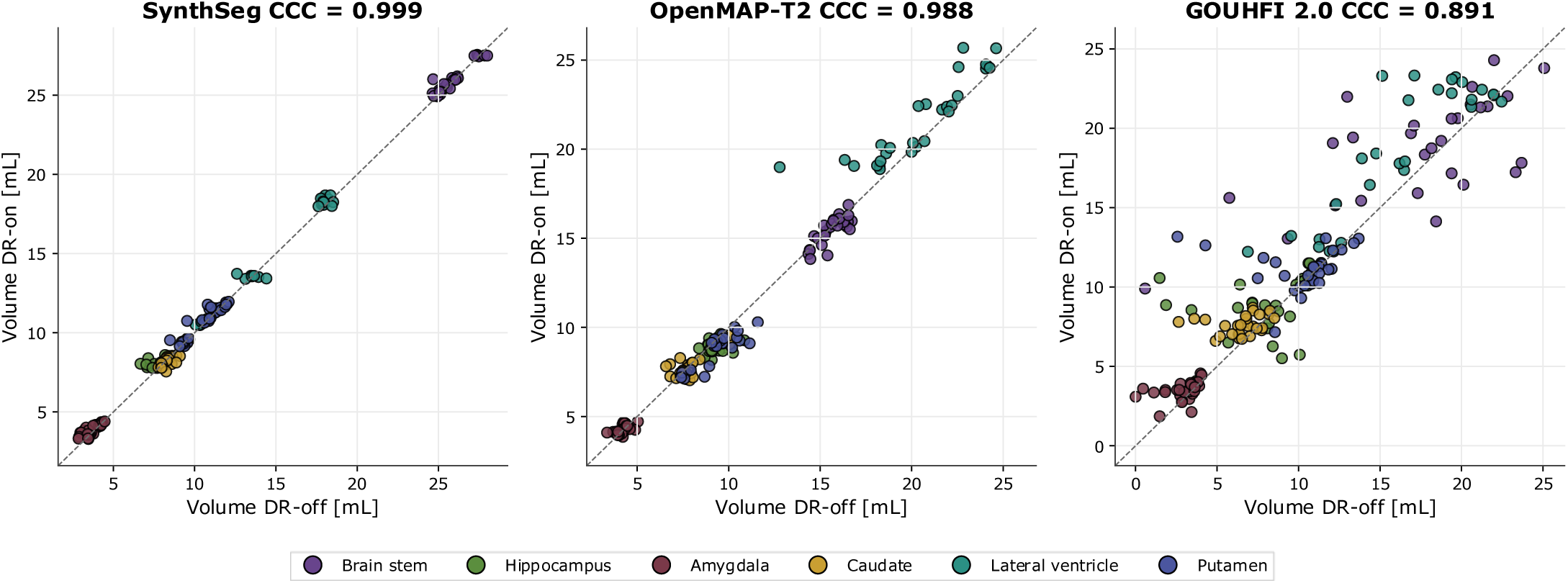
Volume agreement between paired DR-off and DR-on reconstructions per parcellation tool. Each point represents a single subject × acquisition × structure pair (n = 168 per tool; for GOUHFI this includes one DR-off amygdala acquisition whose empty mask is retained as zero volume). The x-axis shows the tool-derived volume under DR-off reconstruction, the y-axis under DR-on reconstruction of the same nominal protocol; the dashed line is the identity y = x. Point colour indicates the anatomical structure (see legend). Lin’s concordance correlation coefficient (CCC) between paired DR-off and DR-on volumes is reported per tool.

### 3.3 Cross-protocol volumetric dispersion by tool and reconstruction state

Cross-protocol dispersion was quantified using the coefficient of variation (CoV) of tool-derived volumes across the seven acquisition protocols within each subject, ROI, tool, and Deep Resolve (DR) reconstruction state. Across all six pipelines, the median CoV was lower for DR-on than for DR-off reconstructions (Table 3). The reduction was particularly pronounced for the three parcellation tools, for which the median CoV decreased from 2.62% to 1.39% for SynthSeg, from 4.42% to 2.14% for OpenMAP-T2, and from 21.75% to 7.87% for GOUHFI. Overall, 88-92% of subject-by-ROI trajectories showed lower CoV values under DR-on than under DR-off reconstruction. Consistent reductions were also observed at the subject level: all subjects exhibited a lower median CoV with DR- on for each of the three parcellation tools, with subject-level CoV ratios (DR-on/DR-off) ranging from 0.30 to 0.57 for GOUHFI, 0.37 to 0.64 for OpenMAP-T2, and 0.44 to 0.58 for SynthSeg.

**Table 3.** Cross-protocol volumetric robustness. CoV computed per (subject × ROI) trajectory across seven protocols; units improved, trajectories with CoV_on < CoV_off.

| Tool | ROI set | CoV DR-off, median | CoV DR-on, median | CoV ratio, median | Subject-median CoV ratio, range |
| --- | --- | --- | --- | --- | --- |
| **SynthSeg** | 18 ROIs | 2.62 | 1.39 | 0.51 | [0.44, 0.58] |
| **OpenMAP-T2** | 19 ROIs | 4.42 | 2.14 | 0.48 | [0.37, 0.64] |
| **GOUHFI** | 19 ROIs | 21.75 | 7.87 | 0.47 | [0.30, 0.57] |
| **TotalSegmentator** | whole-brain | 0.39 | 0.32 | 0.83 | [0.57, 1.08] |
| **HD-BET** | whole-brain | 0.54 | 0.39 | 0.66 | [0.58, 0.97] |
| **NV-Segment-CTMR** | whole-brain | 1.29 | 0.39 | 0.36 | [0.25, 0.50] |

For the three whole-brain tools, cross-protocol dispersion was already low under DR-off reconstruction, with median CoV values ranging from 0.39% to 1.29%, but decreased further under DR-on reconstruction. The largest relative reduction was observed for NV-Segment-CTMR, for which the median CoV decreased from 1.29% to 0.39% (paired CoV ratio, 0.36), with reductions observed in all four subjects. More modest decreases were observed for TotalSegmentator, from 0.39% to 0.32%, and HD-BET, from 0.54% to 0.39%. In absolute terms, the greatest reduction in dispersion was observed for GOUHFI, which also exhibited the highest variability under DR- off reconstruction. Among the whole-brain tools, NV-Segment-CTMR showed the largest relative reduction and the highest baseline cross-protocol variability.

The reduction in cross-protocol dispersion was observed across the full range of measured structure volumes. As illustrated in Figure 2A, DR-on measurements (purple) consistently exhibited lower dispersion than DR-off measurements (green) for all three parcellation tools, encompassing both small subcortical structures and larger cortical regions. These findings indicate that the stabilizing effect of DR-on reconstruction was not restricted to structures of a particular size.

**Figure 2.**
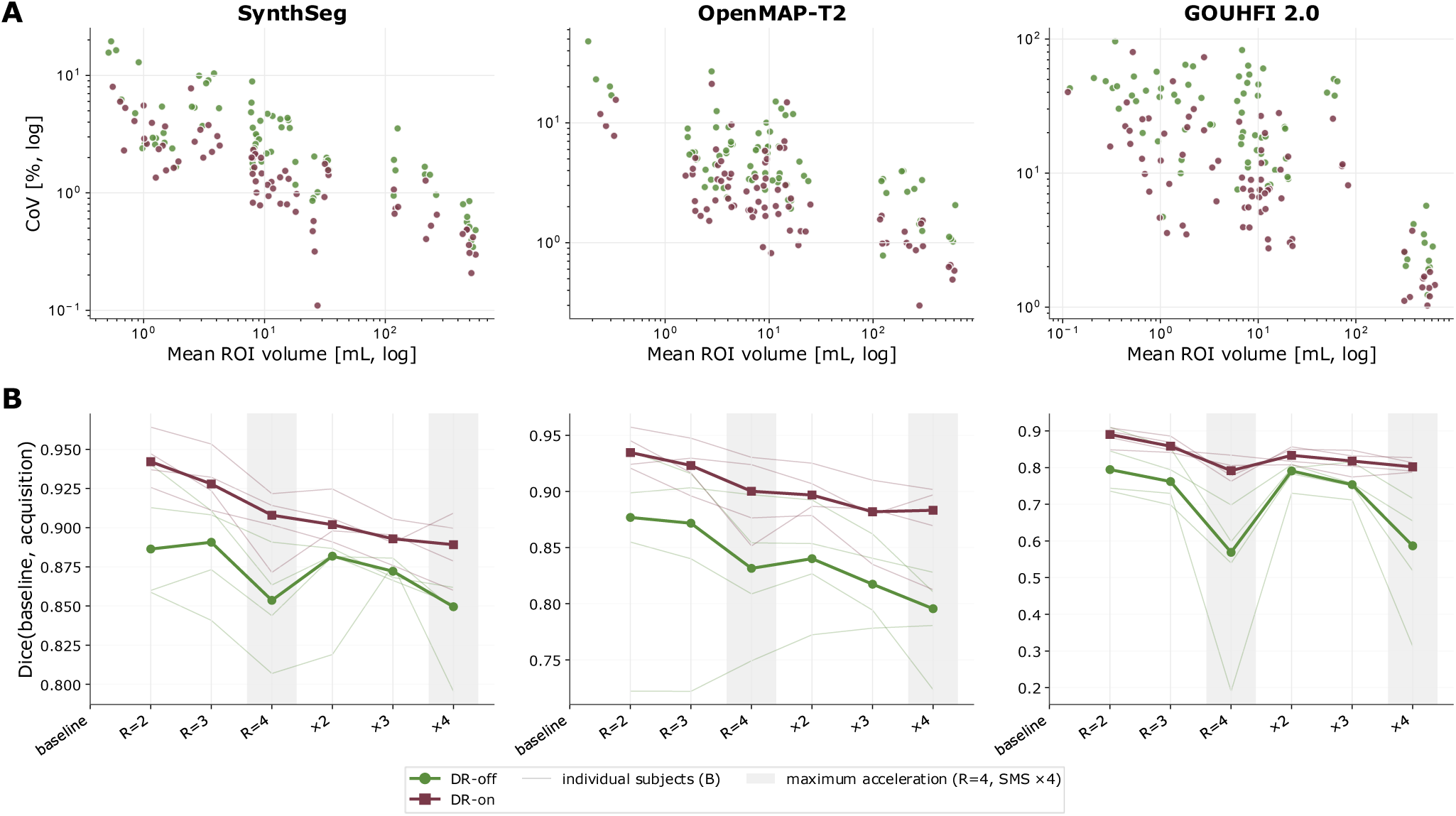
Cross-protocol reproducibility of the three parcellation tools under DR-off (green) and DR-on (purple). (A) Volumetric dispersion: the coefficient of variation of each (subject × ROI) volume trajectory across the seven protocols as a function of the mean ROI volume (log–log). (B) Spatial agreement: the Dice coefficient between each acceleration and the same-Deep-Resolve baseline, across acquisition protocols. Thin lines are individual subjects (each the median over the six manual-reference structures); the bold line is the median across the four subjects. Shaded bands mark the maximum-acceleration protocols (GRAPPA R=4, SMS ×4).

To complement the volumetric coefficient of variation (Figure 2A, Table 3), which quantifies the dispersion of regional volumes across the seven protocols, we evaluated the spatial reproducibility of the parcellations directly. Because all acquisitions of a given subject shared a common geometric frame, label maps were compared without registration, using affine resampling to the baseline grid alone, so that any disagreement reflected the segmentation itself rather than image misalignment. Agreement was quantified with the Dice similarity coefficient, capturing volumetric overlap, and with the 95th-percentile symmetric surface distance (HD95).

Averaged over all pairwise comparisons among the seven protocols, Dice was consistently higher for Deep Resolve reconstructions in every tool (median over subjects and structures). It increased from 0.896 under DR-off to 0.925 under DR-on for SynthSeg, from 0.870 to 0.913 for OpenMAP-T2, and from 0.740 to 0.840 for GOUHFI 2.0. The median minimum pairwise Dice across subject-structure units increased from 0.544 to 0.769 for GOUHFI; the single worst unit fell to a Dice of 0.000 under DR-off (a complete failure) versus 0.512 under DR-on. When each acceleration was referenced to the subject’s own baseline (Figure 2B; each subject summarised by its median over the six structures), Deep Resolve preserved substantially greater overlap as acceleration increased. At GRAPPA R=4 and SMS ×4, respectively, the across-subject median Dice under DR-off was 0.854 and 0.850 for SynthSeg, 0.832 and 0.796 for OpenMAP-T2, and 0.569 and 0.587 for GOUHFI, whereas under DR-on it was 0.908 and 0.889, 0.900 and 0.883, and 0.792 and 0.802. The individual-subject trajectories (Figure 2B) further show that the large DR-off variability of GOUHFI at maximum acceleration - one subject falling well below the others - is strongly attenuated under DR-on.

The surface-distance metric exhibited the same behaviour. Pooled across acceleration levels and structures, the median HD95 relative to baseline decreased under Deep Resolve from 1.03 to 0.51 mm for SynthSeg, from 1.15 to 0.62 mm for OpenMAP-T2, and from 2.89 to 1.54 mm for GOUHFI. The reduction was most pronounced for GOUHFI at maximum acceleration, where HD95 under DR-off and DR-on was 6.00 and 2.43 mm at GRAPPA R=4 and 5.24 and 2.41 mm at SMS ×4; the already small distances for SynthSeg and OpenMAP-T2 tightened further, from 1.09 to 0.73 mm and from 1.34 to 0.77 mm at GRAPPA R=4, respectively. Because the acquisitions comprised 6 mm through-plane slices, HD95 chiefly reflects in-plane boundary agreement, with through-plane disagreements registering in coarse, slice-sized increments.

### 3.4 Volume changes across all acceleration levels

Per-acquisition Deep Resolve associated volume changes for the three parcellation tools are reported for all seven protocols in Table 4. For SynthSeg and OpenMAP-T2, pooled median ΔV% was within ±1.5% at baseline and at moderate acceleration (R = 2 or 3, SMS ×2 or ×3), rising to +2.9% and −6.7%, respectively, at maximum acceleration. For GOUHFI, ΔV% was already appreciable at intermediate acceleration and reached +31.51% at GRAPPA R = 4 and +31.98% at SMS ×4.

**Table 4.** Per-acquisition Deep Resolve associated volume change (ΔV%) across all seven protocols, pooled across subject × structure pairs. Median [IQR]; n = 24 pairs per acquisition per tool.

| Acquisition | SynthSeg $\Delta V\%$ | OpenMAP-T2 $\Delta V\%$ | GOUHFI $\Delta V\%$ |
| --- | --- | --- | --- |
| baseline | -0.27 [-1.10, +0.65] | +0.41 [-1.85, +2.15] | +0.88 [-4.06, +3.06] |
| GRAPPA R=2 | +0.18 [-0.92, +0.92] | -0.30 [-2.03, +1.29] | +2.64 [-1.22, +5.34] |
| GRAPPA R=3 | +0.68 [-0.87, +1.74] | -0.60 [-3.00, +1.83] | +4.35 [-2.15, +14.8] |
| GRAPPA R=4 | +2.11 [+0.21, +11.7] | -6.71 [-12.6, -0.51] | +31.51 [+15.59, +60.18] |
| SMS $\times 2$ | +0.19 [-0.13, +1.34] | +0.14 [-1.29, +0.90] | +2.57 [-4.75, +6.51] |
| SMS $\times 3$ | +0.72 [-0.45, +1.82] | -1.32 [-4.09, +1.20] | +10.34 [+0.40, +15.3] |
| SMS $\times 4$ | +2.91 [-0.39, +7.30] | -2.21 [-8.66, +11.5] | +31.98 [+21.6, +68.7] |

Whole-brain tools (TotalSegmentator, HD-BET, NV-Segment-CTMR) subject-level median $\Delta V\%$ remained within $\pm 1.5\%$ at every acquisition level;

To characterise acceleration-related volume changes separately within each Deep Resolve state, deviations relative to the corresponding baseline acquisition were computed at maximum acceleration as δ = 100 · (V_acq / V_baseline - 1) for the six manual-reference structures. For SynthSeg and OpenMAP-T2, DR-off produced moderate deviations (median |δ| 6.6 to 7.2%) that DR-on reduced to 1.4 to 4.3%. No SynthSeg or OpenMAP-T2 pair produced a volume below half of the same-Deep Resolve baseline in either state. For GOUHFI, DR-off produced systematic under-segmentation (median δ = −31.6% at GRAPPA R = 4, −25.6% at SMS ×4), 7 of 24 pairs at R = 4 and SMS ×4 falling below half of the same-Deep Resolve baseline. Under DR-on reconstruction, GOUHFI median |δ| decreased to 8.1% at R = 4 and 5.1% at SMS ×4, and no pair fell below the failure threshold. The 14 GOUHFI failure events observed under DR-off reconstruction occurred in three of the four subjects and involved five of the six structures.

Figure 3 illustrates these patterns. For SynthSeg and OpenMAP-T2, the DR-off and DR-on median deviations and subject ranges largely overlapped and remained close to zero. In contrast, for GOUHFI, DR-off median deviations were negative for most ROIs at both GRAPPA R = 4 and SMS ×4, with subject ranges extending toward larger negative values. Under DR-on reconstruction, the deviations were generally closer to zero and the subject ranges were narrower. Figure 4 shows the corresponding volume trajectories across all seven protocols. For GOUHFI, DR- off median volumes and individual-subject trajectories decreased at the two highest acceleration settings across the evaluated structures, with the largest reductions observed for the lateral ventricle and hippocampus. Under DR-on reconstruction, the trajectories generally remained closer to their corresponding baseline volumes. Figure 5 illustrates this qualitatively for the putamen: under DR-off the GOUHFI segmentation fragments at GRAPPA R = 4 and SMS ×4, whereas DR-on restores a complete structure closely matching the manual STAPLE reference, while SynthSeg and OpenMAP-T2 remain stable throughout.

**Figure 3.**
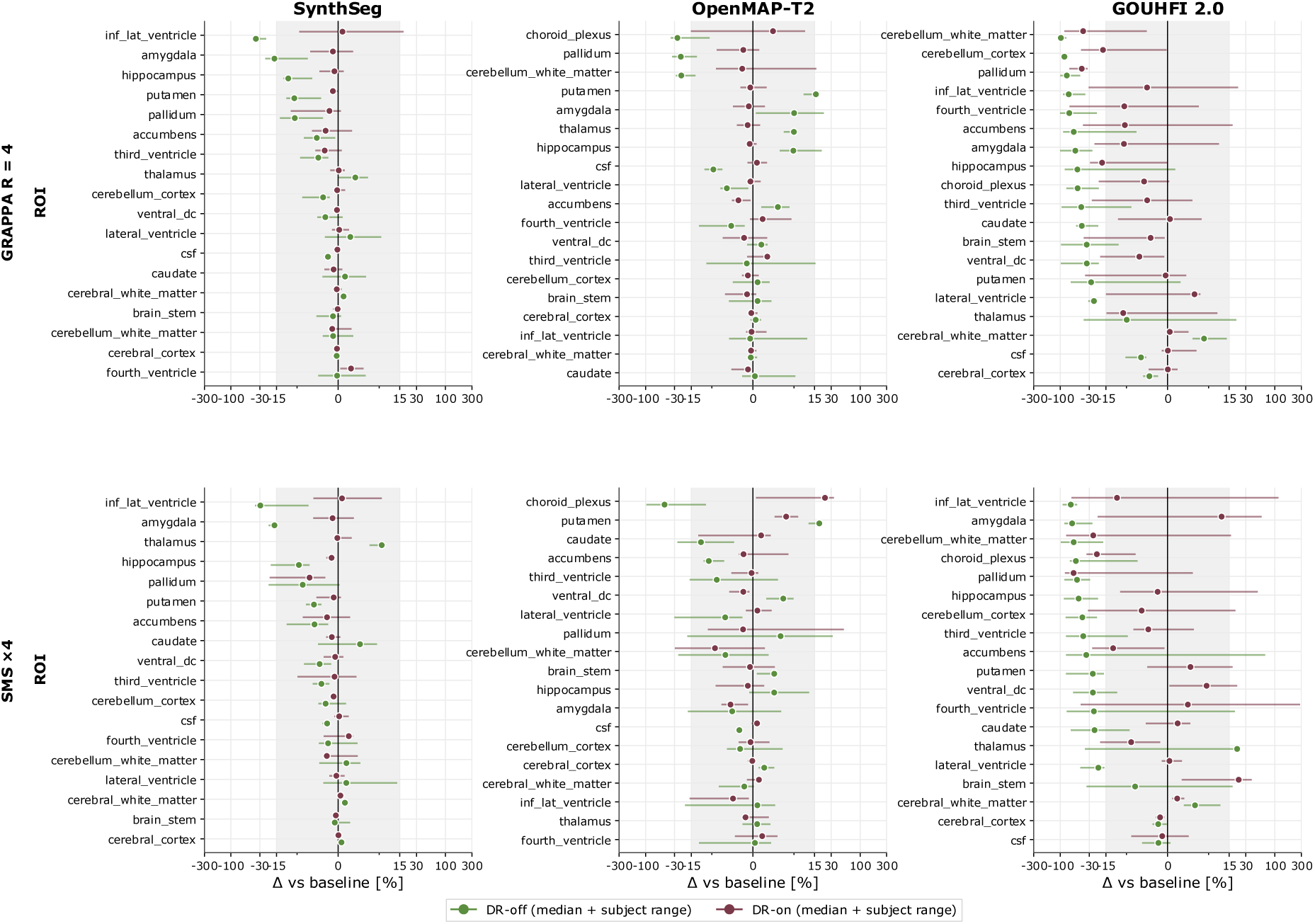
Volume deviation from the same-Deep Resolve baseline at maximum acceleration for all available parcellation ROIs. For each (subject × ROI × tool × Deep Resolve state) combination, deviation δ = 100 · (V_acq / V_baseline − 1) is computed with V_baseline taken from the baseline acquisition of the matching Deep Resolve state. Within each panel, dots mark the subject-level median across four subjects and horizontal whiskers span the subject minimum-to-maximum range; green = DR-off, purple = DR-on.

**Figure 4.**
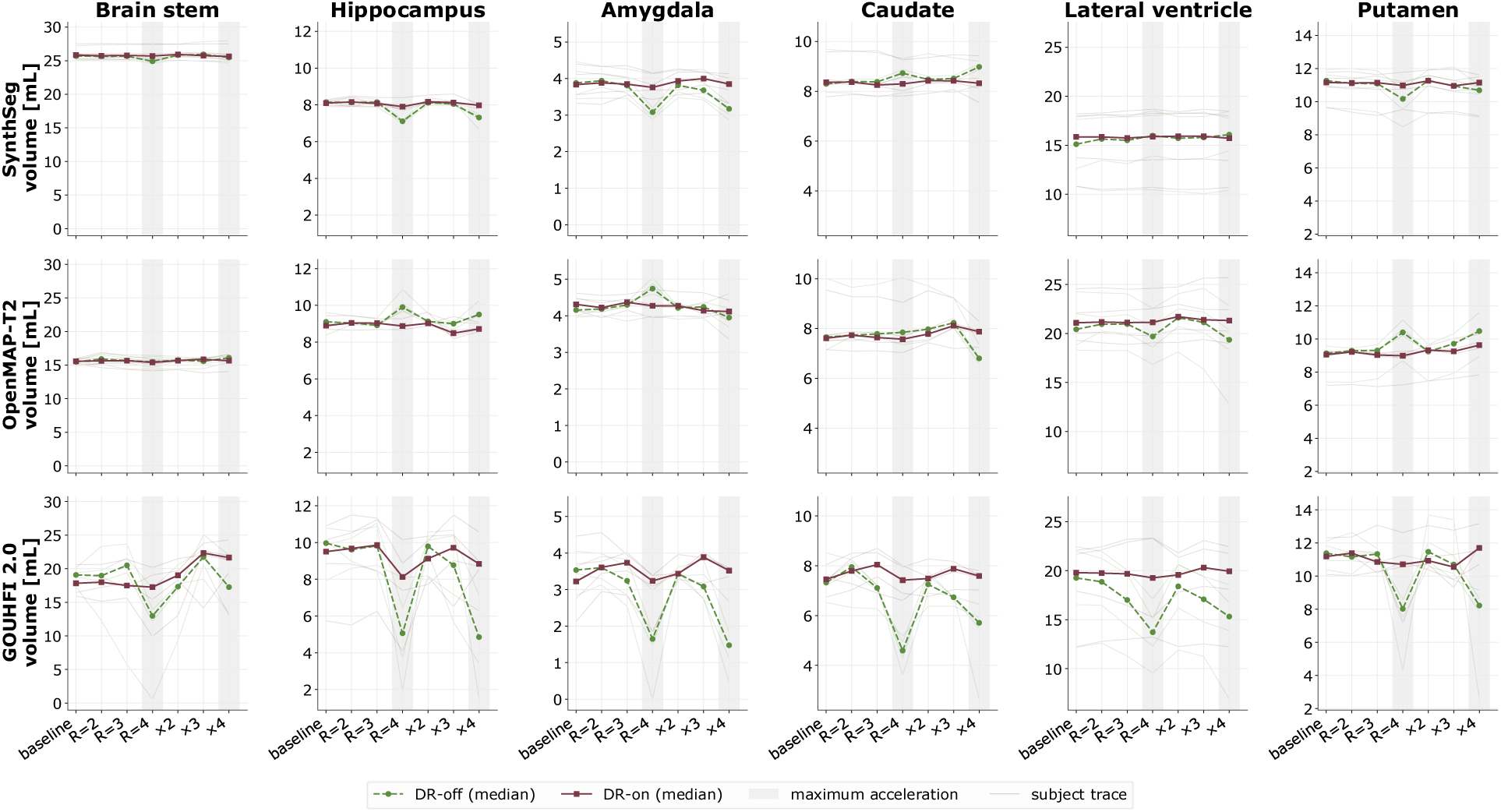
Volume trajectories across the seven acquisition protocols for the six manual-reference structures. Within each panel, thin lines are individual subject trajectories (four per Deep Resolve state) and bold lines are the median across subjects; dashed green = DR-off, purple = DR-on. The shaded grey bands mark the two maximum-acceleration protocols (GRAPPA R = 4, SMS ×4).

**Figure 5.**
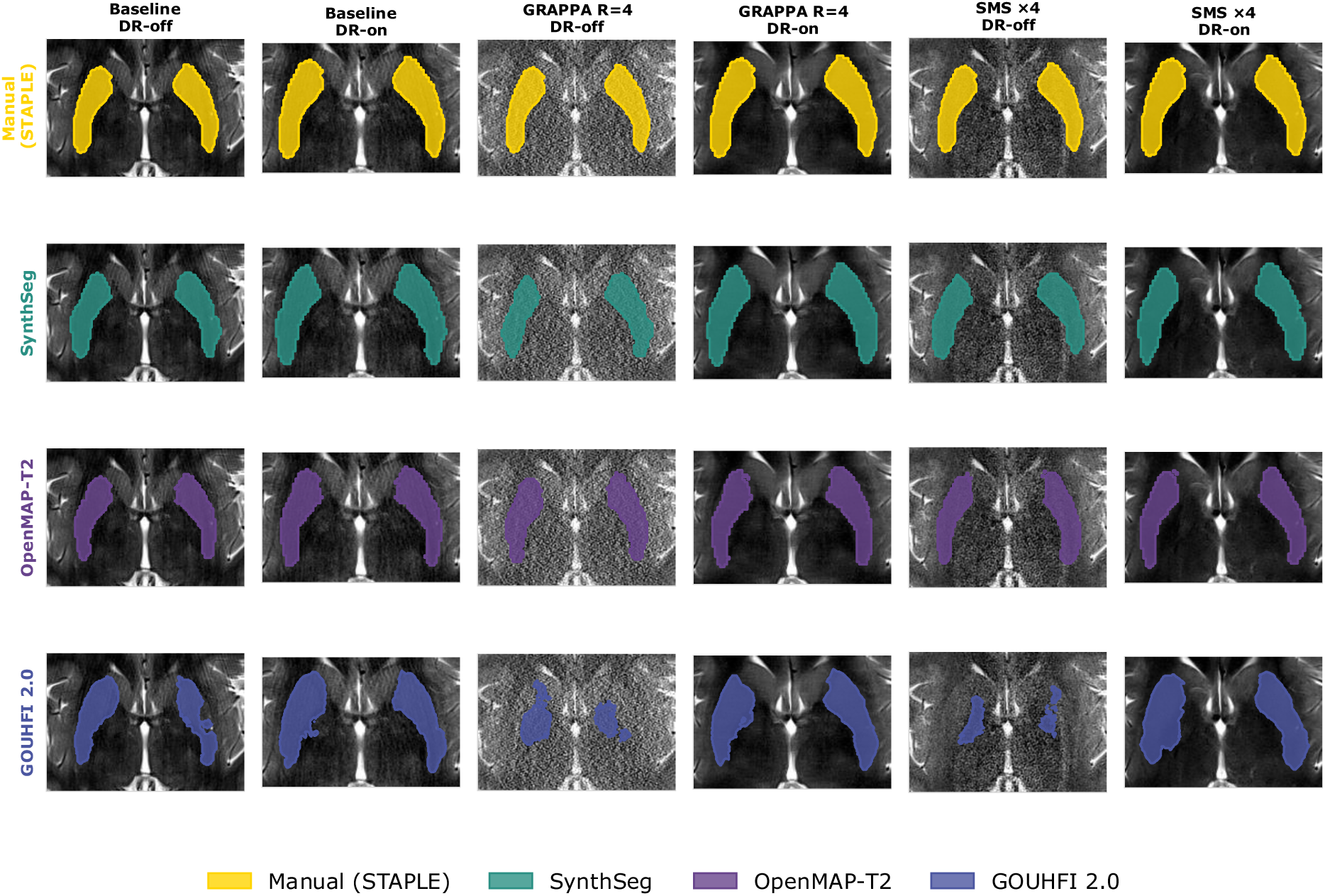
Putamen segmentation across acquisitions for a representative subject. Rows correspond to the segmentation source - the manual STAPLE consensus and the three parcellation tools (SynthSeg, OpenMAP-T2, GOUHFI 2.0) - and columns to the acquisition: baseline, GRAPPA R = 4, and SMS ×4, each in the DR-off and DR-on reconstruction state.

### 3.5 Whole-brain volume stability across protocols and reconstruction states

Whole-brain volume estimates obtained with the three whole-brain segmentation tools were less sensitive to acquisition protocol and Deep Resolve (DR) reconstruction state than the regional volume estimates derived from the parcellation tools (Table 5). TotalSegmentator and HD-BET showed high stability under DR-off reconstruction, with median paired ΔV% values across subjects of +0.19% and −0.33%, respectively, and corresponding cross-protocol CoV values of 0.39% and 0.54%. The additional reduction in variability associated with DR-on reconstruction was therefore relatively modest for these two tools, with CoV decreasing by 18% for TotalSegmentator and 28% for HD-BET. In contrast, NV-Segment-CTMR exhibited greater subject- and protocol-dependent variability under DR-off reconstruction, with a median paired ΔV% of +0.93% across subjects (IQR, +0.14% to +1.27%) and a cross-protocol CoV of 1.29%. Under DR-on reconstruction, its CoV decreased to 0.39%, corresponding to an approximately two-thirds reduction in cross-protocol variability. Deviations from the same-Deep Resolve baseline at R = 4 and SMS ×4 remained within ±2% for all three whole-brain tools in both reconstruction states. Figure 6 confirms these patterns visually: subject-level whole-brain volumes differ systematically between tools by tens of millilitres (Panel A), individual subject clusters lie very close to the identity line for TotalSegmentator and HD-BET while NV-Segment-CTMR shows wider scatter driven by one subject (Panel B), and per-subject trajectories are essentially flat across the seven protocols with DR-off and DR-on lines overlapping tightly (Panel C). Because no manual whole-brain reference was available in this study, whole-brain results are reported as concordance and stability rather than accuracy.

**Figure 6.**
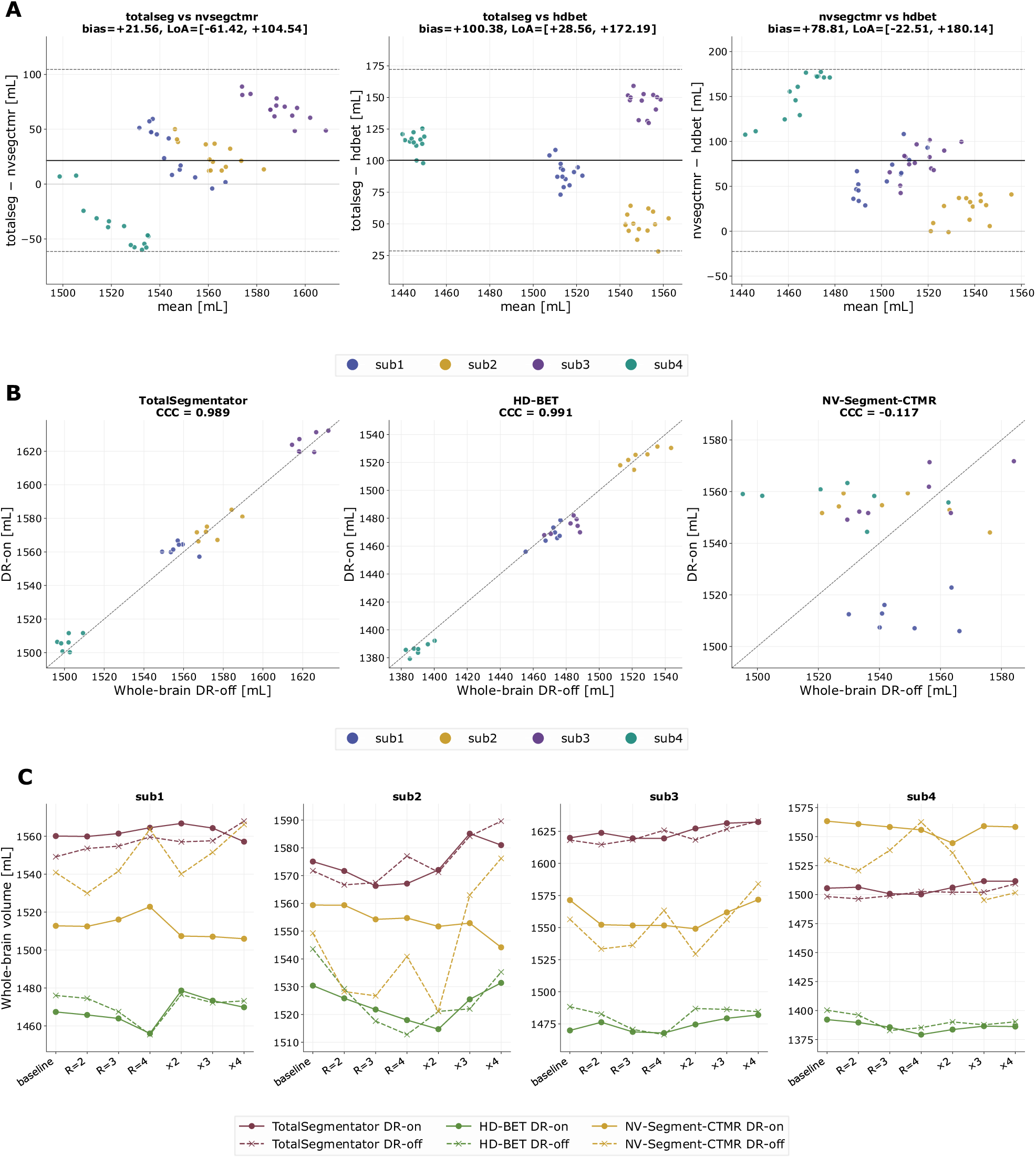
Whole-brain volume stability across protocols and Deep Resolve states for the three brain-extraction pipelines. Panel A: Bland–Altman plots of the pairwise whole-brain volume difference between the three brain-extraction tools across all 56 acquisitions. Bias (mean paired difference) and 95% limits of agreement (LoA, dashed grey) are annotated per pair. Panel B: whole-brain volume under DR-off (x-axis) versus DR-on (y-axis) reconstruction of the same nominal protocol per tool (n = 28 pairs per tool). Dashed line is the identity y = x. Lin’s concordance correlation coefficient (CCC) between paired DR-off and DR- on volumes is reported per tool. Panel C: whole-brain volume trajectory across the seven acquisition protocols, shown separately for each subject (four columns). Solid marker/line = DR-on, dashed marker/line = DR-off;

**Table 5.** Whole-brain volume stability. ΔV%, paired difference DR-on minus DR-off across 28 pairs; δ, median deviation from same-Deep Resolve baseline across 4 subjects.

| Tool | $\Delta V\%$, median [IQR] | | CoV, % | | GRAPPA R = 4, $\delta$ % | | SMS $\times 4$, $\delta$ % | |
| --- | --- | --- | --- | --- | --- | --- | --- | --- |
|  | Paired | Subject-level | DR-off | DR-on | DR-off | DR-on | DR-off | DR-on |
| **TotalSegmentator** | +0.24 [+0.03, +0.44] | +0.19 [+0.09, +0.31] | 0.39 | 0.32 | +0.41 | -0.19 | +1.02 | +0.39 |
| **HD-BET** | -0.25 [-0.47, +0.05] | -0.33 [-0.43, -0.23] | 0.54 | 0.39 | -1.43 | -0.79 | -0.40 | +0.12 |
| **NV-Segment-CTMR** | +0.60 [-1.28, +1.42] | +0.93 [+0.14, +1.27] | 1.29 | 0.39 | +0.96 | -0.39 | +1.69 | -0.38 |

### 3.6 Resolution-control analysis

To assess whether the previously observed Deep Resolve–associated differences in volumetric stability could be attributed to the change in reconstructed in-plane resolution, a resolution-control analysis was performed using SynthSeg as a representative parcellation pipeline. Figure 7 shows normalized volume trajectories across all seven acquisition protocols for native DR-off and DR-on reconstructions together with their corresponding resolution-matched controls.

**Figure 7.**
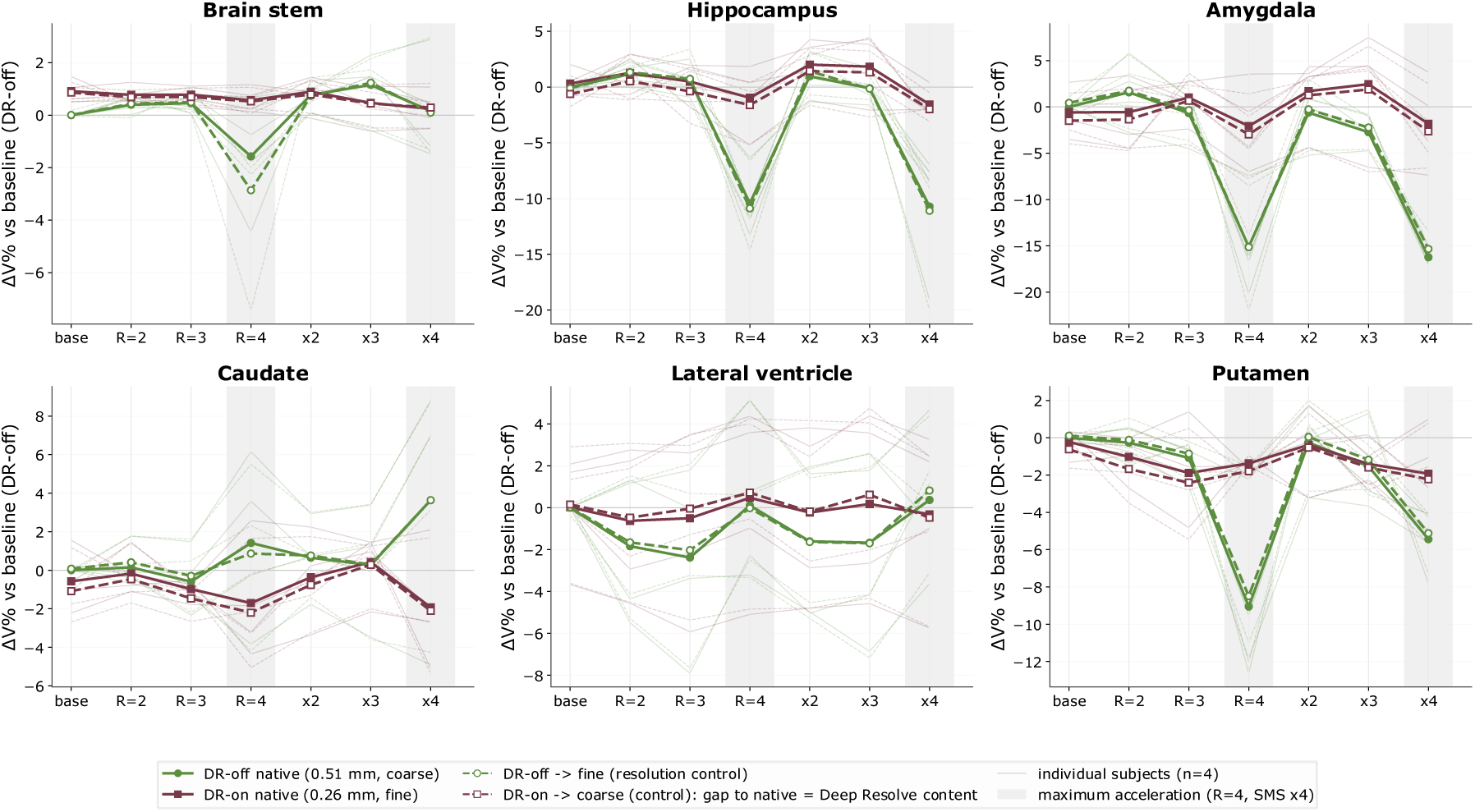
Resolution-control analysis of SynthSeg-derived regional volumes across acquisition protocols. Green lines indicate DR-off and purple lines DR-on. Solid lines represent native-resolution images, and dashed lines with open markers the corresponding resolution-matched controls. Thin lines show individual participants (n = 4), and thick lines show the mean across participants.

Across the six evaluated structures, the resolution-matched controls closely followed the native trajectories within each Deep Resolve state. Increasing the spatial sampling of DR-off images to match the nominal in-plane resolution of DR-on did not eliminate the volume deviations observed at higher acceleration factors. Conversely, reducing the spatial resolution of DR-on images to that of DR-off did not substantially alter the DR-on trajectories. This pattern was most apparent at GRAPPA R = 4 and SMS ×4, where the separation between DR-off and DR-on measurements increased for several structures, particularly the hippocampus, amygdala, and putamen, while the corresponding native and resolution-controlled trajectories within each reconstruction state remained closely aligned.

Quantitative decomposition of the observed volume differences further supported this finding. The median absolute contribution attributable to resolution remained small and nearly constant across acquisition protocols, ranging approximately from 0.16% to 0.29%. In contrast, the reconstruction-related component increased with acceleration and was most pronounced at the highest acceleration settings. At GRAPPA R = 4, the median absolute resolution and reconstruction components were 0.29% and 3.49%, respectively, corresponding to a resolution contribution of approximately 8%. At SMS ×4, the respective values were 0.27% and 5.20%, with resolution accounting for approximately 5% of the combined effect magnitude.

Overall, the resolution-control analysis indicates that differences in reconstructed in-plane resolution accounted for only a minor proportion of the Deep Resolve–associated volumetric effect. The persistence of DR-off and DR- on differences after resolution matching, together with the increasing reconstruction-related component at higher acceleration factors, supports the interpretation that the improved volumetric stability observed with DR- on primarily reflects reconstruction-dependent image characteristics rather than the change in spatial sampling alone.

## 4. Discussion

This study examined how sensitive automated brain-volume measurements are to the parallel-imaging acceleration and to deep-learning reconstruction on prospectively acquired T2-TSE MRI, across three subcortical parcellation pipelines and three whole-brain tools, benchmarked against a three-rater STAPLE consensus. Three findings stand out. First, Deep Resolve improved reference-relative accuracy most for GOUHFI 2.0 (pooled median absolute percentage error 28.39% to 21.89%), modestly but consistently for SynthSeg (16.19% to 15.53%), and had a mixed, structure-dependent effect for OpenMAP-T2. Second, and more consistently, Deep Resolve substantially reduced cross-protocol dispersion: median coefficient of variation approximately halved for all three parcellation tools (SynthSeg 2.62% to 1.39%, OpenMAP-T2 4.42% to 2.14%, GOUHFI 21.75% to 7.87%), with 88 to 92% of subject-by-ROI trajectories improving, and spatial reproducibility improved in parallel (mean pairwise Dice rising and median HD95 relative to baseline falling for every tool). Third, Deep Resolve eliminated the systematic GOUHFI under-segmentation seen at high acceleration under conventional reconstruction: DR-off produced a median deviation of −31.6% at GRAPPA R = 4 and 14 failure events falling below half of the same-reconstruction baseline, spanning three of four subjects and five of six structures, whereas no such failure remained under DR-on. Whole-brain masking was robust in both states, with already-low dispersion that decreased further, the largest relative reduction being for NV-Segment-CTMR (median coefficient of variation 1.29% to 0.39%).

### 4.1 Clinical significance

These patterns matter precisely because the clinical questions that drive automated volumetry turn on small differences. Hippocampal and whole-brain atrophy rates used to support a diagnosis of neurodegeneration, or to detect progression, are of the same order as the cross-protocol variability observed here under conventional reconstruction: annualised hippocampal atrophy in Alzheimer’s disease averages approximately 4.7%, against approximately 1.4% in age-matched controls [1], a separation comparable in magnitude to the several-percent measurement shifts that acceleration and reconstruction can themselves introduce. A pipeline whose output shifts by several percent when a site changes its acceleration factor, or when a scanner software update alters the default reconstruction, can generate apparent longitudinal change where there is none, or mask real change. The dispersion reduction afforded by Deep Resolve is therefore of direct clinical value: not because it makes any single measurement more accurate, but because it makes serial and multi-protocol measurements more comparable. The lateral ventricle, the structure showing both the largest reference-relative error and the largest Deep Resolve benefit (paired median change in error of −8.35 pp for GOUHFI and −3.20 pp for OpenMAP-T2), is instructive here, as ventricular volume is a widely used surrogate for parenchymal loss in demyelinating disease, where the brain parenchymal fraction and ventricular enlargement track neurodegeneration and long-term disability independently of focal lesion burden [2], and for cerebrospinal-fluid-space enlargement in hydrocephalus, where automated ventricular volumetry is increasingly used for diagnosis and shunt follow-up [3].

### 4.2 Reconstruction as a source of quantitative bias

A deeper implication concerns the nature of deep-learning reconstruction itself. Modules such as Deep Resolve Gain and Sharp do not merely denoise; through adaptive denoising and learned super-resolution they actively reshape image content, and this reshaping propagates into any downstream measurement that depends on tissue boundaries. Consistent with this, the DR-off and DR-on differences observed here persisted after resolution matching and grew with acceleration factor, indicating that the improved stability reflects reconstruction-dependent image characteristics rather than the change in spatial sampling alone. The two-sided nature of this effect is visible in the data: the same operation that suppresses acceleration-induced degradation can also shift absolute values, as with GOUHFI, whose Deep-Resolve-associated volume change reached +31.5% at GRAPPA R = 4. This is consistent with prior reports that accelerated acquisition biases automated volumetry [8] and that deep-learning reconstruction preserves quantitative performance only partially [14, 15]. The practical consequence is that reconstruction state must be treated as a fixed factor in any quantitative protocol: mixing DR-off and DR-on data within a longitudinal series, or across sites, risks a systematic bias that downstream statistical harmonisation cannot fully recover.

The same reshaping affects other quantitative image descriptors. On comparable GRAPPA- and SMS-accelerated acquisitions from the same scanner platform, Deep Resolve has been shown to alter the stability of radiomic texture features in a tissue- and acceleration-dependent manner [16], so the volumetric effect reported here is one facet of a broader influence of deep-learning reconstruction on image-derived measurements. This matters clinically because such descriptors increasingly serve as decision-support tools: radiomic texture analysis can flag occult lesions that are not readily apparent on visual inspection, for example periapical lesions on intraoral radiographs [24]. Where feature values underpin an automated decision, an image-formation change that shifts those values could alter the decision itself. The principle is therefore not specific to one vendor, to the brain, or to T2 weighting: comparable deep-learning reconstruction pipelines are now deployed across musculoskeletal, abdominal, and cardiac imaging, and the message, that image-formation algorithms are an active variable in quantitative imaging and must be reported and controlled as such, generalises across anatomy and modality.

### 4.3 Tool dependence and pipeline selection

The order-of-magnitude difference in robustness between pipelines has immediate practical weight. Whole-brain extraction was essentially protocol-agnostic, so applications requiring only global parenchymal volume can tolerate heterogeneous acquisition. Fine-grained subcortical parcellation was far more fragile, and the most sensitive pipeline (GOUHFI) was also the one most rescued by reconstruction, meaning that pipeline choice and reconstruction state interact and cannot be selected independently. For clinical or multi-site research deployment, this argues for validating a specific segmentation pipeline against the specific acquisition and reconstruction settings in use, rather than assuming that a tool validated on T1-weighted or on unaccelerated data will behave equivalently on accelerated, deep-learning-reconstructed T2-weighted images.

### 4.4 Limitations

Several limitations temper these conclusions. The sample comprised four healthy adult volunteers scanned on a single 1.5 T system with one 2D T2-TSE protocol family at 6 mm slice thickness; generalisation to pathological anatomy, to other field strengths and vendors, to 3D acquisitions, and to thinner sections cannot be assumed. The expert reference was a STAPLE consensus of three raters on the DR-off baseline acquisition, which estimates rather than establishes ground truth; absolute percentage error therefore captures volumetric agreement and is insensitive to spatially compensating segmentation errors. Because the acquisitions used 6 mm through-plane slices, the surface-distance metric chiefly reflects in-plane boundary agreement, with through-plane disagreements registering in coarse, slice-sized increments. Finally, the study is cross-sectional; the longitudinal reproducibility that most directly determines clinical utility was inferred from cross-protocol dispersion rather than measured over repeated sessions. Prospective evaluation in patient cohorts, across vendors, and with true longitudinal repeat imaging is the natural next step.

## 5. Conclusions

On prospectively acquired T2-TSE brain MRI, deep-learning reconstruction (Siemens Deep Resolve) had a limited-to-positive effect on the absolute accuracy of automated volumetry, largest for GOUHFI 2.0, and consistently improved its stability across acceleration protocols, approximately halving cross-protocol variability for subcortical parcellation, improving spatial reproducibility, and eliminating the systematic under-segmentation that the most acceleration-sensitive pipeline exhibited at high acceleration factors. Robustness was strongly pipeline- and structure-dependent: whole-brain masking was protocol-agnostic, whereas fine subcortical parcellation required reconstruction support to remain reliable. Because clinically meaningful volumetric change is often of the same magnitude as protocol-induced variability, reconstruction state should be treated as a controlled, reported variable in any quantitative imaging protocol, and segmentation pipelines should be validated under the acquisition and reconstruction conditions in which they will be used. Although demonstrated here in the brain, these considerations apply to any setting in which deep-learning image formation precedes automated quantification.

## 6. Declarations

## Author contributions

Conceptualization, R.O. and M.S.; methodology, R.O., M.S., J.L. and A.K.; MRI acquisition and data curation, R.O., I.D., W.K., I.O. and J.M.; software and formal analysis, J.L. and M.S.; investigation, R.O., J.L. and W.K.; visualization, J.L. and M.S.; writing—original draft preparation, R.O., J.L. and M.S.; writing—review and editing, all authors; supervision, M.S. and R.O. All authors have read and agreed to the published version of the manuscript.

## Ethics approval and consent to participate

The study protocol was designed in accordance with guidelines of the Declaration of Helsinki and the Good Clinical Practice Declaration Statement. Written approval was obtained from the Bioethics Committee to conduct this study (No. 11/KBL/OIL/2025 dated 11 March, 2025 and 03 March 2026). Written informed consent was obtained from all volunteers prior to scanning.

## Consent for publication

Not applicable; the manuscript contains no individually identifiable participant data.

## Data availability

The analysis and figure-generation code, together with the manuscript figures, are publicly available on Zenodo: https://doi.org/10.5281/zenodo.22664823. The source MRI images and segmentation masks are available from the corresponding author on reasonable request.

## Use of generative AI and AI-assisted technologies

During the preparation of this manuscript, the authors used generative AI tools to assist with language editing, structural organization, and literature-search support. All AI-assisted text and bibliographic details were critically reviewed and verified by the authors, who take full responsibility for the content of the final manuscript.

